# Reducing visible aerosol generation during phacoemulsification in the era of Covid-19

**DOI:** 10.1101/2020.05.14.20102301

**Authors:** Kieren Darcy, Omar Elhaddad, Asaf Achiron, Johannes Keller, Duncan Leadbetter, Derek Tole, Sidath Liyanage

## Abstract

**Objective:** To assess potential methods of reducing visible aerosol generation during clear corneal phacoemulsification surgery in the era of Covid-19.

**Methods:** Aerosol generation during phacoemulsification was assessed using a model comprising a human cadaveric corneoscleral rim mounted on an artificial anterior chamber. Typical phacoemulsification settings were used and visible aerosol production was recorded using high speed 4K camera. Aerosolisation was evaluated under various experimental settings: Two different phacoemulsification tip sizes (2.2mm, 2.75mm), varying levels of corneal moisture, the use of suction and blowing air in the surgical field, the use of hydroxypropyl methylcellulose (HPMC) coating of the cornea with a static and moving tip.

**Results:** This model demonstrates visible aerosol generation during phacoemulsification with a 2.75mm phacoemulsification tip. No visible aerosol was noted with a 2.2mm tip. The presence of visible aerosol is unrelated to corneal wetting. Suction in close proximity to the aerosol plume did not impact on its dispersion. Blowing air redirected the aerosol plume towards the ocular surface. Visible aerosol production was abolished when HPMC was used to coat the cornea. This effect lasted for an average of 67±8 seconds in the static model. Visible aerosol generation was discerned during movement of the 2.2mm tip towards the corneal wound.

**Conclusions:** We demonstrate visible aerosol production in the setting of a model of a clear cornea phacoemulsification. Visible aerosol can be reduced using a 2.2mm phacoemulsification tip and reapplying HPMC every minute during phacoemulsification.

## Introduction

Covid-19 is a pandemic disease with presentations that range from asymptomatic cases, to mild respiratory infection, to severe acute respiratory syndrome with associated mortality (1). The causative organism (coronavirus SARS-CoV-2) is transmitted via fomites, droplets and aerosols carrying infective particles usually originating from an infected person’s respiratory tract (2–4). There is concern that healthcare workers may contract Covid-19 during aerosol generating procedures (AGP) performed on patients carrying the disease (5). This has been shown to be of particular importance in bronchoscopy, endotracheal intubation and extubation (3).

Phacoemulsification surgery is one of the most common operations performed worldwide (6). Some ophthalmologists have concerns that it may be an AGP based on the high-speed oscillations used to generate ultrasonic energy and the aerosol or ‘vapour’ plume that is sometimes observed during energy delivery. The detection of SARS-CoV-2 genetic material in conjunctival swabs in a small proportion of affected patients has raised the possibility of transmission of Covid-19 disease by this aerosol (7,8). It is unknown how long it will take for the pandemic to be controlled, whether and when effective vaccinations or treatments will be developed, and how routine medical care may be performed in a manner that is safe for patients and healthcare workers. The purpose of this paper is to evaluate different methods of reducing visible aerosol generation during phacoemulsification surgery.

## Methods

### Setting

This study was performed in accordance with the Declaration of Helsinki. A fresh human cadaveric corneoscleral rim was obtained from the NHS Blood and Transplant for the training and research purposes. The rim was fitted on a Coronet™ artificial anterior chamber (AAC) (Network Medical, Ripon, UK) and inflated with balanced salt solution (BSS; Balanced salt solution; Beaver-Visitec International, Oxford, UK). This was mounted within a model head used for cataract training (Phillips Studio, Bristol, UK). A triplanar clear cornea incision was performed for use with sleeved 45-degree angled Kelman phacoemulsification tips (Centurion Vision System; Alcon, Camberley, UK). Typical phacoemulsification settings were used (continuous torsional energy (90%), intra-ocular pressure 60 mmHg, vacuum 475mmHg, aspiration flow 33cc/min, continuous irrigation on). Aerosol generation was assessed using high speed 4K camera (Canon 5D mark IV, 100mm prime macro lens; Canon, Tokyo, Japan) and lighting (Rotolight Neo II; Rotolight, Iver Heath, UK) under various experimental settings:

A. Different sizes of phacoemulsification tips Two phacoemulsification tips (2.2mm 0.9mm Intrepid FMS, 2.75mm 0.9mm ABS; Alcon, Camberley, Surrey) were used with matching clear corneal wounds (ClearCut; Alcon, Camberley, Surrey).
B. Dry/wet status of the cornea The cornea was wetted by application of BSS on the surface. Drying of the corneal surface was achieved using cellulose eye spears (Weck-Cel, Beaver-Visitec International, Oxford, UK).
C. Suction and blowing air Suction was achieved using a standard Yankauer suction tip at maximal suction and a Bair Hugger™ (3M, Minnesota, USA) was used to blow ambient-temperature air in the vicinity of the surgical field.
D. Hydroxypropyl methylcellulose 2% on the corneal surface The use of hydroxypropyl methylcellulose 2% (HPMC) (Aurovisc; Aurolab, Madurai, India & Beaver-Visitec International, Oxford, UK) on the corneal surface was compared to BSS. The duration between the reduction in aerosolisation with the application of HPMC and the observation of aerosol was measured 6 sequential times. Copious BSS irrigation was used to clear the residual HPMC between each trial.
E. Static/dynamic setting The static setting was achieved by mounting the phacoemulsification probe on a vertical stand for fixation and the tip was introduced 2.5mm (using a reference mark on the sleeve) towards the centre of the AAC reproducing standard orientation (Fig. 1). In the dynamic trial, the surgeon handled the phacoemulsification probe to move the tip inside the AAC to simulate nuclear sculpting, longitudinally along its axis.

**Fig. 1.**
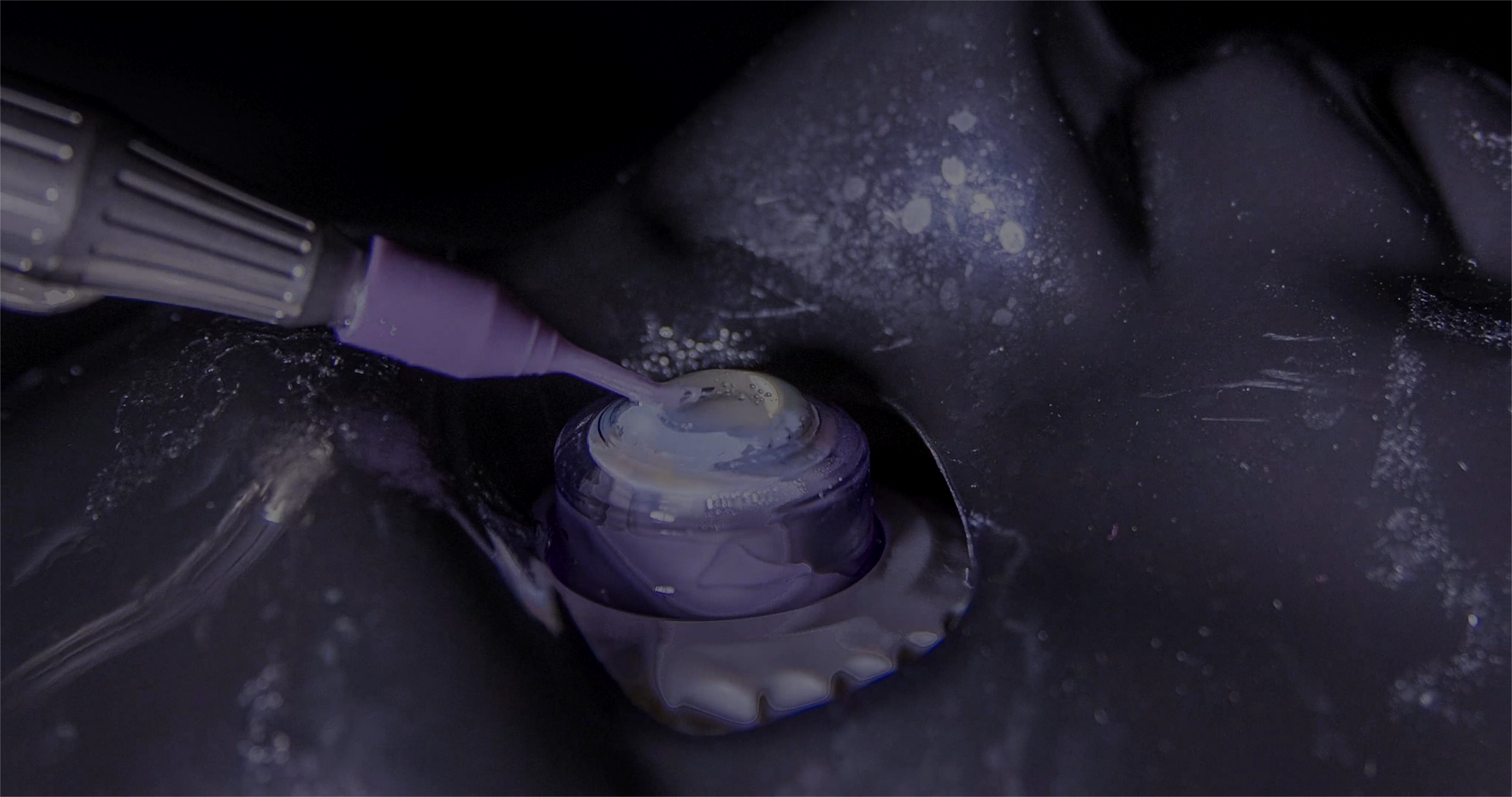
Photograph showing the corneoscleral rim mounted on the anterior artificial chamber. This is sited within a model head used for cataract training. The phacoemulsification probe is fixed in position.

## Results

This model used a human cadaveric corneoscleral rim and an artificial anterior chamber to demonstrate aerosol production during phacoemulsification. Phacoemulsification was performed using a standard 2.75mm phacoemulsification tip with typical ultrasound and phacofluidic settings (Fig. 2, Video 1). No visible aerosol was identified when using the 2.2mm tip with identical ultrasound and phacofluidic settings (Fig. 3, Video 2).

**Fig. 2.**
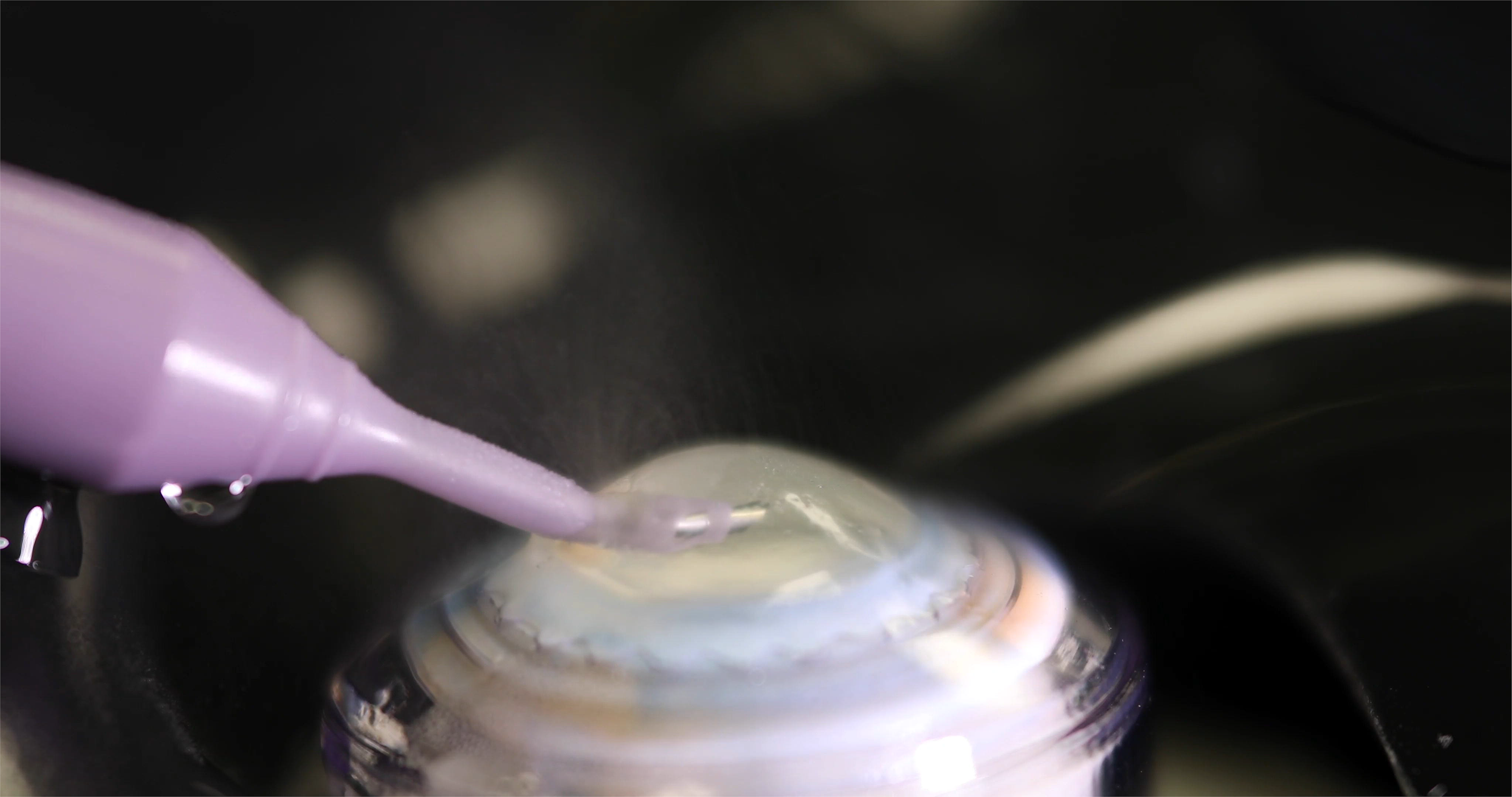
Photograph showing visible aerosol produced using continuous torsional phacoemulsification with the 2.75mm tip

**Fig. 3.**
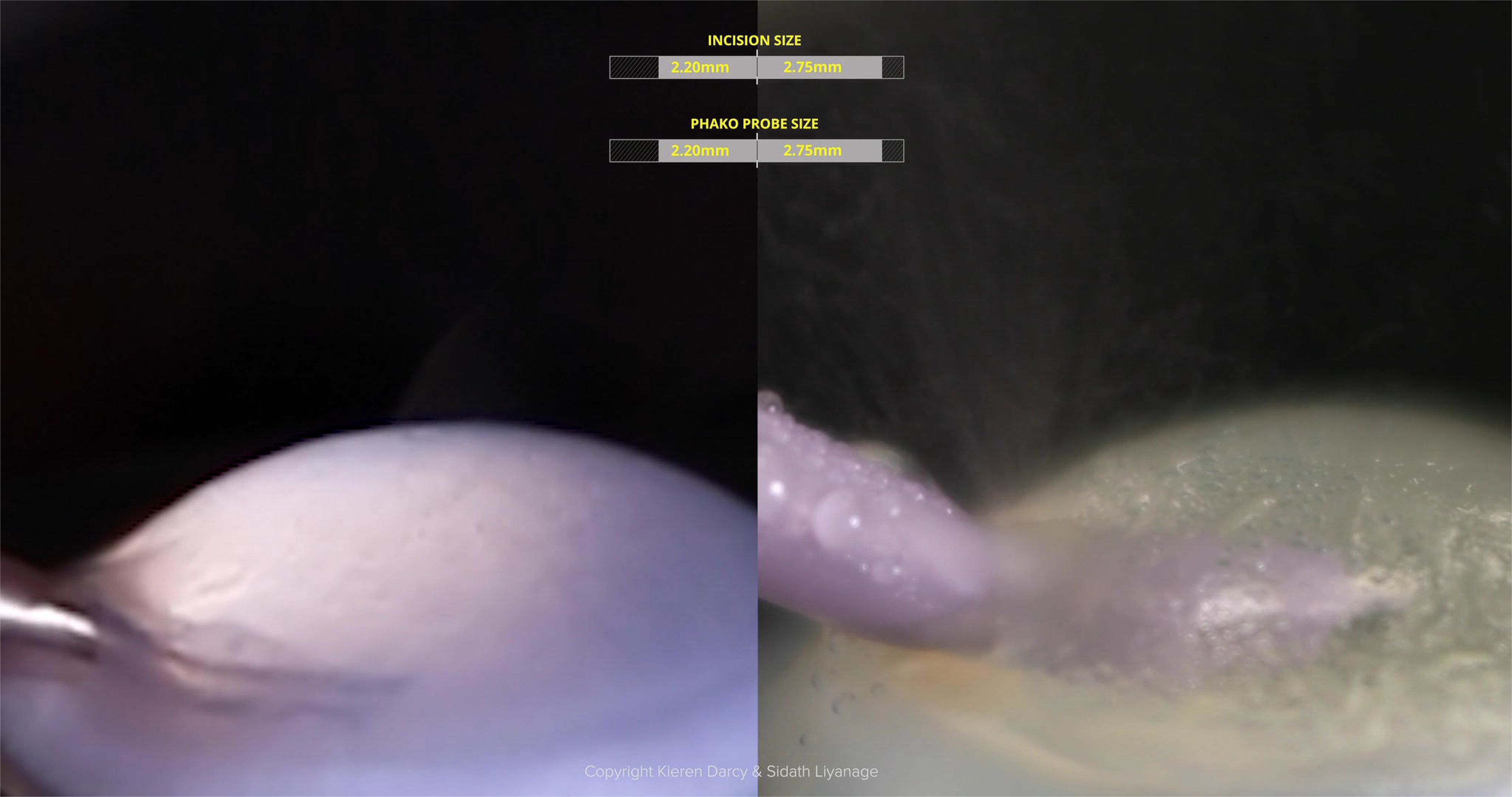
Photograph comparing visible aerosol production with the 2.2mm and 2.75mm tip

The aerosol produced by the 2.75mm tip did not vary depending on corneal wetting. This aerosol was still evident when the corneal surface was dried (Video 3). Attempts to aspirate the aerosol plume generated by negative pressure or suction using Yankauer suction were unsuccessful (Fig. 4a, Video 4). Downward positive air pressure using forced air at ambient temperature altered the morphology of the plume generated, redirecting the upward aerosol plume downwards and then laterally along the ocular surface (Fig. 4b, Video 5).

**Fig. 4.**
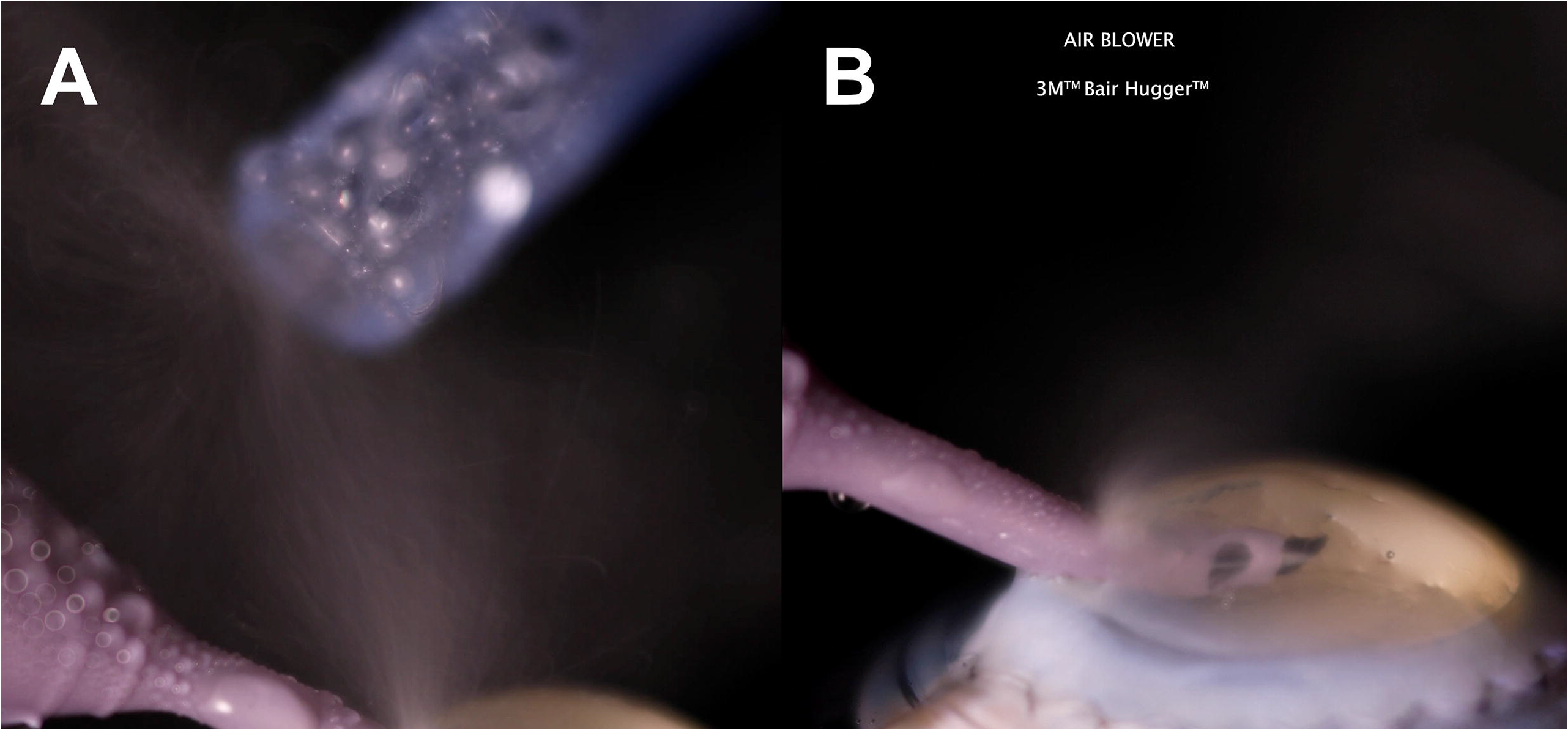
Photographs showing (A) failure to aspirate visible aerosol using maximal Yankauer suction and (B) redirection of aerosol plume downwards by blowing air

Coating the corneal surface with HPMC resulted in a prominent change: no visible aerosol production was demonstrated during phacoemulsification in the static model where the handpiece was fixed within the AAC (Fig.5, Video 6). This effect lasted for an average of 67±8 seconds (range 63–72s) of continuous phacoemulsification in the static model (Video 7).

**Fig. 5.**
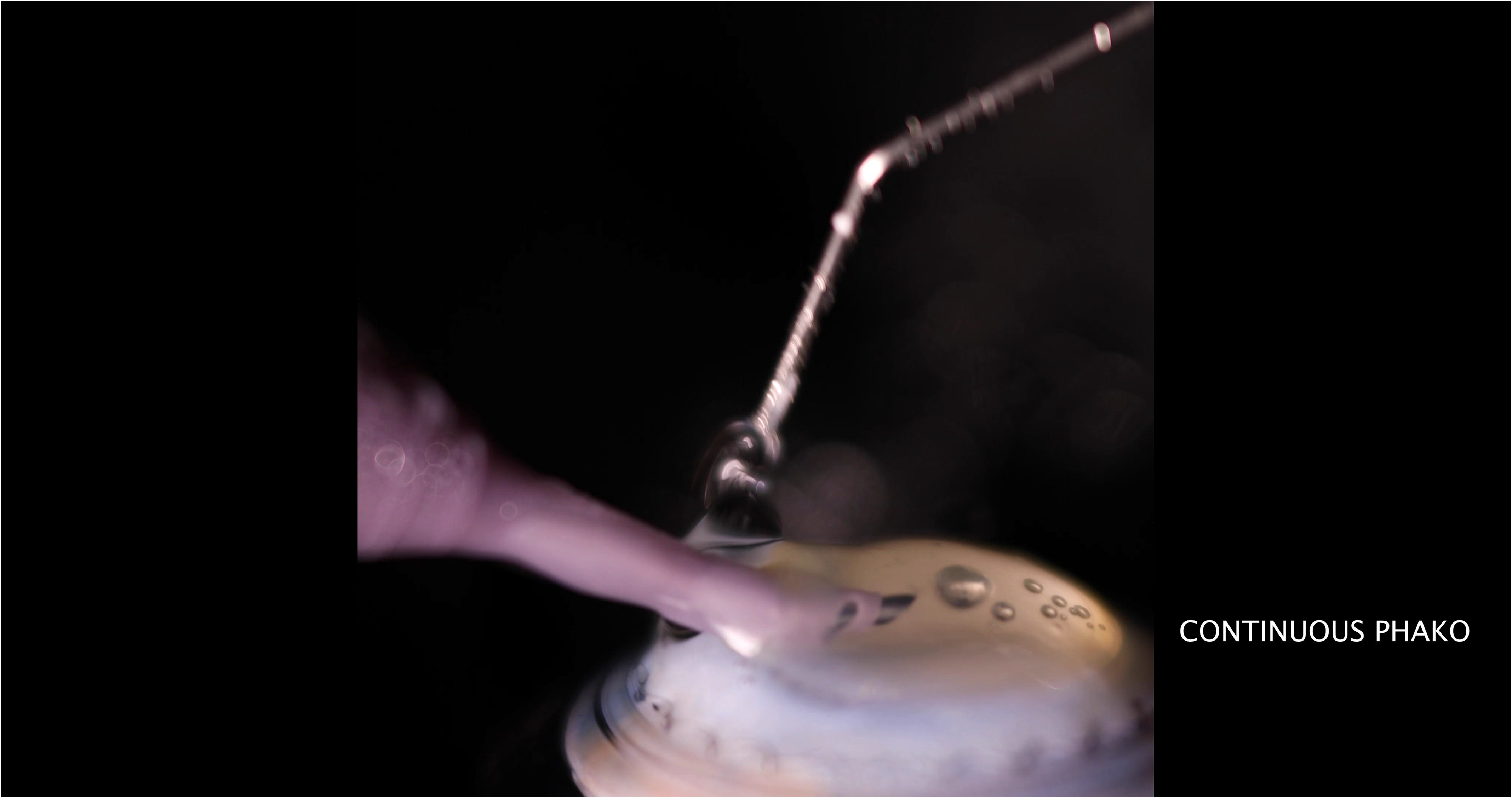
Photograph showing the cessation of visible aerosol during phacoemulsification on application of HPMC

When continuous phacoemulsification was performed in a dynamic fashion, with the 2.2mm tip mimicking the longitudinal action of nuclear sculpting, no visible aerosol generation was discerned during the majority of the movement. The exception was noted when the tip was just proximal to the corneal wound when the aerosol plume was clearly visualised (Video 8).

## Discussion

Since the advent of the Covid-19 pandemic, the risk to healthcare workers contracting the disease from potentially infectious droplets and aerosol during aerosol generating procedures (AGPs) has been explored by several studies analysing the pattern of droplet spread during simulated medical interventions (9,10).

Simulations published online have attempted to model aerosol generation during phacoemulsification using video recording and application of the ultrasound energy at either an air-fluid interface or using model eyes (11). The former is not representative of the closed system that is required for effective and safe phacoemulsification. Similarly, plastic model eyes have different structural properties to the human eye and due to this, it is challenging to extrapolate any meaningful conclusions using these models. Currently, there are no published reports of similar studies in the peer-reviewed literature.

Artificial anterior chambers (AACs) are utilised in corneal graft surgery for donor tissue preparation (12). Once the human corneoscleral rim is mounted on the AAC, a single leak-free chamber is formed allowing the maintenance of ‘intraocular pressure’. This model allows simulated surgery for lamellar graft procedures using human cadaveric tissue and addresses the above limitations of other experimental models.

We utilised a novel application of this model to consistently demonstrate the aerosol or ‘vapour’ plume that is observed in the real world setting during phacoemulsification with a 2.75mm tip. The plume appears to originate from the corneal wound. This may be due to vibration transmitted from the phacoemulsification probe to the lip of the corneal wound. It is impossible to determine whether the aerosol generation seen with this experimental set up is due to agitation of moisture on the surface of the cornea or small amounts of BSS that can egress through the corneal wound around the probe. Visible aerosol following emulsification with a dry cornea supports the latter. The reduction in visible aerosol seen with the 2.2mm tip may be explained by less wound leakage. This was noted in an ex vivo study comparing wound leakage in an ex vivo comparison of 2.2mm and 2.75mm tips (13).

Following this, we evaluated different methods of attenuating this aerosol plume and demonstrate two potential approaches. The first approach involves forced air directed at the microsurgical field to disperse the aerosol downwards and laterally. In a operating theatre, this set up would be challenging as the air would have to be filtered and humidified to prevent infection and corneal drying respectively with the setup of the microsurgical field to incorporate a suitable air flow device potentially being problematic. Portable surgical laminar flow systems used for cataract surgery may be configured to drive any aerosol produced away from healthcare workers, allowing precipitation onto surgical drapes and subsequent disposal.

Suction did not influence the plume despite close proximity of the Yankauer suction tip to its source and maximum suction. This is likely to be due to a limitation in the instrument used, as custom devices have been developed to effectively remove vapour during corneal ablation using excimer laser (14).

The second approach uses hydroxypropyl methylcellulose 2% (HPMC) to coat the corneal surface during phacoemulsification. HPMC is a low-viscosity and moderate viscoelasticity material commonly used to coat the cornea to improve intraocular visualisation during surgery and has proven safety (15).

Analysis of video recordings highlight that visible aerosol generation is abolished when HPMC is applied to the corneal wound. This effect lasted for approximately one minute. It is possible that when HPMC coats the corneal wound and phacoemulsification probe, it acts as a barrier to either aerosol formation or dispersion. Its higher molecular weight and elasticity may also act as a damper to the ultrasonic energy preventing agitation and consequent aerosolisation. This effect was consistent using two different brands of HPMC and may apply to other types of viscoelastic materials.

Efforts to reproduce the longitudinal action of nuclear sculpting showed that, in contrast to the static model, visible aerosol was only produced when continuous phacoemulsification was performed close to the corneal wound despite HPMC coating. The close proximity of the ultrasound energy to the cornea may generate aerosol by either agitating the HPMC coat off the cornea or directly diminishing HPMC’s barrier function. The reduced aerosol production at other points during the dynamic experiment may be due to less deformation of the corneal wound by the surgeon despite efforts to minimise this when using the static model and represents a limitation of this study. This study only analysed aerosol production using continuous torsional phacoemulsification and may not be representative of other modes of ultrasound energy delivery.

This study investigated visible aerosol generation from phacoemulsification using a clear corneal wound and may not represent aerosol produced from a limbal corneal wound. As this study focused on visible aerosol generation only, it is impossible to comment on generation of invisible aerosol particles that are not discernable by this experimental setup. There is uncertainty about whether these particles can form in the absence of visible aerosol generation and if so, whether these particles pose a risk of transmitting Covid-19 disease to healthcare workers.

The first part of this study aimed to reproduce the ‘vapour’ plumes observed in real world cataract surgery. To the best of our knowledge, this study is the first in peer-reviewed published literature to demonstrate visible aerosol generation during phacoemulsification.

We cannot comment on the risk of transmission from an infected patient to healthcare workers by the visible aerosol generated. Currently, it is unknown whether viable SARS-CoV-2 particles can be aerosolised in sufficient quantities to form an effective inoculum for transmission to healthcare workers. The following should be considered when estimating this risk: is there an effective SARS-CoV-2 viral load present in or on the ocular tissues of an asymptomatic or undiagnosed patient undergoing cataract surgery, and is the visible aerosol plume formed during cataract surgery is composed of BSS or intraocular fluids. We offer suggestions to deal with these uncertainties.

Topical povidone-iodine is virucidal with in vitro testing of 0.23% povidone-iodine producing a 10,000-fold reduction in the viral load of SARS-CoV and MERS-CoV within 15 seconds (16–18). Topical povidone-iodine is widely used in ophthalmic surgery for surgical antisepsis and would destroy any viable SARS-CoV-2 particles present on the ocular surface.

There may be concern that the aerosol generated by phacoemulsification could contain virus-infected intraocular fluid. The majority of this intraocular fluid is visco-expressed before capsulorrhexis. The concern about any remaining infected intraocular fluid should be alleviated by applying statistical and mathematical principles. The mean volume of the anterior chamber measured with optical coherence tomography is 0.17±0.04 ml (19). If the volume of the anterior chamber is normally distributed, three standard deviations would 99.7% of the population would have an anterior chamber volume of less than 0.29 ml.

Phacoemulsification cataract surgery occurs in a closed system and irrigating/aspirating BSS through the anterior chamber with the phacoemulsification probe results in a dilution of intraocular fluid that obeys first order decay: for every 0.29 ml of BSS that passes through, the concentration of the original intraocular fluid is decreased by 50%. When 2.03 ml of BSS has circulated in the anterior chamber, the concentration is reduced to less than 1% of the original concentration. Using conventional aspiration rates ranging from 20–40 cc/min, this will take at most 3 to 6 seconds in 99.7% of the population. Performing this irrigation/aspiration step for a aspiration rate-dependent duration prior to phacoemulsification will significantly decrease the proportion of any remaining intraocular fluid (and consequently any theoretical virus load) in any aerosol produced.

We then investigated methods of mitigating of any potential risk from the aerosol produced by phacoemulsification, with the prospect of aiding the resumption of elective cataract surgery as we navigate through this pandemic. Findings from the study show that use of a 2.2mm phacoemulsification tip where possible and copious administration of HPMC every minute during phacoemulsification reduces visible aerosol generation.

## Data Availability

All available data is included in manuscript.

## Acknowledgements

We wish to acknowledge the support of NHS Blood and Transplant.

